# Utilization of Emergency Care by an East African Refugee Community Compared to Local Residents in Central Minnesota: A Cross-Sectional Study

**DOI:** 10.1101/2022.09.30.22280583

**Authors:** Abby Hughes-Scalise, Mohamed Ayanle, Srikanth Dakoji

**Author notes:** <u>Corresponding author:</u> Abby Hughes-Scalise, PhD, LP, 2211 Riverside Ave, Minneapolis, MN 55454.

## Abstract

**Background:** Previous studies on emergency service utilization by refugees have been mixed, with some showing overuse and others showing underuse relative to host populations. Much of the previous literature focuses on refugees by continent of origin, which may mask important differences within immigrant groups. Limited research has investigated emergency medicine utilization for East African refugees in the United States.

**Methods:** The current study investigated differences in ED service utilization for East African refugees compared to local residents at an Emergency Trauma Center (Level 2) in Central Minnesota. From a convenience sample of 48 East African refugees and 116 local residents that presented to an emergency department (ED) in Central Minnesota, survey data was collected on self-reported reasons for presenting to ED; chart review data was collected on care received at the emergency department and discharge diagnoses.

**Results:** Logistic regressions showed that refugees were significantly (p < .05) more likely than local residents to self-report difficulties with navigating the primary care settings as a reason for presenting to the ED, and less likely to report needing specialized care. Refugees were also significantly less likely than local residents to call their primary care provider/clinic before presenting to the ED. During the ED visit, refugees were significantly more likely than local residents to receive symptomatic care and less likely to receive advanced imaging.

**Conclusion:** We hope these results spur additional research on this unique population, particularly related to health literacy and health equity in an emergency medicine setting for East African refugees continuing to migrate to the United States at high rates.

## INTRODUCTION

Over the past 2 decades, there has been a steady rise in the numbers of people fleeing their homeland due to persecution, violence, human rights violations, and climate change. Just in the first half of 2021, more than 89.3 million individuals were forcibly displaced worldwide —the highest number in the past 2 decades.^[1]^ In 2019, Sub-Saharan African immigrants accounted for 5% of all refugees currently residing in the United States, and 35% of Sub-Saharan immigrants came from East African countries.^[2]^ According to the most recent United States census data, East African refugees from Somalia make up the second-largest group of immigrants residing in Minnesota, with more than 33,000 East African refugees and over 58,000 Minnesota residents reporting Somali ancestry.^[3]^

The health and wellbeing of refugees is a public health crisis met with many challenges, exacerbated by differences in access to health care between refugees and host populations.^[4]^ Notable inequities include: lack of provider understanding of refugee health issues, language and cultural barriers, racial discrimination, and higher costs of refugee health care.^[5][6]^ These barriers often lead to unmet health care needs, delayed diagnosis and treatment of health conditions, inability to receive preventative care, financial burdens, and morbidity.^[7][8]^

Important access points to healthcare services are emergency departments (EDs) and primary care clinics. Previous research on healthcare utilization of these services by refugee populations has shown mixed results. Studies outside of the United States suggest that refugees utilize more services overall, and higher ED visit frequencies in particular compared to host populations.^[9]^ However, in a recent US sample, refugees did not differ from local residents in their use of primary care, and refugees utilized ED services less frequently than local residents.^[10]^ Limited research has been conducted on why refugees are accessing services differently, though some findings include: fear of security implications if they were to call for emergency medical help,^[11]^ lack of health literacy,^[6]^ and limited English proficiency.^[12]^

Literature regarding emergency service utilization by East African refugees in the United States is scant. As previous literature on refugee healthcare utilization often groups refugees by continent of origin, or even across continents of origin, a critique of the current literature is that findings about broad refugee populations may mask important differences within immigrant groups.^[13]^ As such, there is a call for more research focused on specific refugee populations.

The current study aims to fill this important gap in the literature by investigating differences in ED service utilization of East African refugees compared to local residents at an Emergency Trauma Center (Level 2) in Central Minnesota. We examined differences in patient-reported reasons for utilizing ED, services received during their visit, and discharge diagnoses.

## METHODS

### Patient and public involvement

Neither patients nor the public were involved in developing the current study’s research questions, design, or recruitment. They will not be involved in our plans to disseminate study results.

### Participants

This cross-sectional study was conducted in an ED, Level 2 Trauma Center, in Central Minnesota that serves a large East African refugee population. Data was collected from a convenience sample of individuals who presented to the ED between 11/24/2014 and 06/09/2015. During this time, approximately 32,000 individuals presented to this ED; 5% of those individuals identified Somali as their primary language.

Refugee patients who presented to the ED meeting all inclusion criteria were invited to participate: refugee participants were required to be of East African descent, to acknowledge that Somali was the primary language spoken at home, and to have immigrated to the United States within the last 5 years. Local resident patients meeting all inclusion criteria were invited to participate: participants were required to have been born in the US and to acknowledge that English was the primary language spoken at home.

Patients with major trauma or mental health crisis who were not able to communicate (e.g., intubated, delirium) were excluded from the current study.

### Ethics Statement

The study was granted approval by the Saint Could Hospital’s Institutional Review Board prior to participant recruitment. Participants were asked to read and sign a consent form, a HIPAA authorization form, and an assent form (if participant was under the age of 18) prior to study participation. If translation was needed, it was provided by either a research assistant who was fluent in Somali or one of the ED’s full-time interpreters. Written consent (and assent, if applicable) was obtained from all participants included in the current study.

### Data Collection

Demographic data collected from chart review included age, gender, country of origin, treatment course during their ED visit, and ED discharge diagnosis. A self-reported survey was also completed by participants during their ED visit, described below.

### Measures

The self-report survey was generated by the third author and gathered data on the following: employment status, insurance coverage, whether participants had an identified primary care provider, and whether participants contacted their primary care provider/clinic prior to their ED visit. The questionnaire also asked participants to specify their reasons for coming to the ED rather than a primary care clinic. The following possible reasons were listed for choosing to come to the ED: (a) no appointment available; (b) clinic is closed; (c) bad experience at clinic; (d) needed specialized care; (e) need to be admitted; (f) family reasons (e.g., no child care during the day); (g) work conflict (e.g., no sick time); (h) no transportation; and (i) other.

To facilitate meaningful analysis, researchers coded the responses into three categories: clinic difficulties, the need for specialized care, and psychosocial reasons. If participants endorsed (a), (b), or (c), they were coded as a ‘yes’ for clinic difficulties. If participants did not endorse all three of those reasons, they were coded as a ‘no’ for clinic difficulties. In a similar fashion, if participants endorsed (d) or (e), they were coded ‘yes’ for needing specialized care. If participants endorsed (f), (g), or (h), they were coded ‘yes’ for psychosocial reasons.

ED treatment course and ED discharge diagnosis were gathered via chart review and further coded by the researchers. ED treatment course was organized into three categories: receiving symptomatic care (e.g., intravenous fluids, oral and/or intramuscular medications), receiving specialty interventions not generally administered in a clinic setting (e.g., cardiac monitoring, administration of intravenous antibiotics, bolus and continuous infusion of antiarrhythmic medications), and receiving advanced imaging (e.g., ultrasound, CT, MRI). All ED treatment course coding was completed by at least two of the study authors. ED diagnosis codes were organized into 14 broad diagnostic categories, as well as an “other” category.

### Statistical Analysis

Descriptive analyses were run for demographic variables in order to contextualize the sample. Binary logistic regressions were performed to ascertain the effects of refugee status on the likelihood that participants would endorse various reasons for presenting to the ED, as well as to determine the effects of refugee status on the likelihood that participants would receive various types of care in the ED. Logistic regressions were also used to predict various discharge diagnoses from refugee status.

## RESULTS

### Demographic Results

During the data collection period, 164 participants consented to participate:116 local residents and 48 refugees of East African descent. Of the 48 refugee participants, 39 identified Somalia as their country of origin. An additional five refugee participants identified Kenya, three refugee participants identified Ethiopia, and one refugee participant identified Tanzania. Participant demographic information is summarized in Table 1.

**Table 1:**
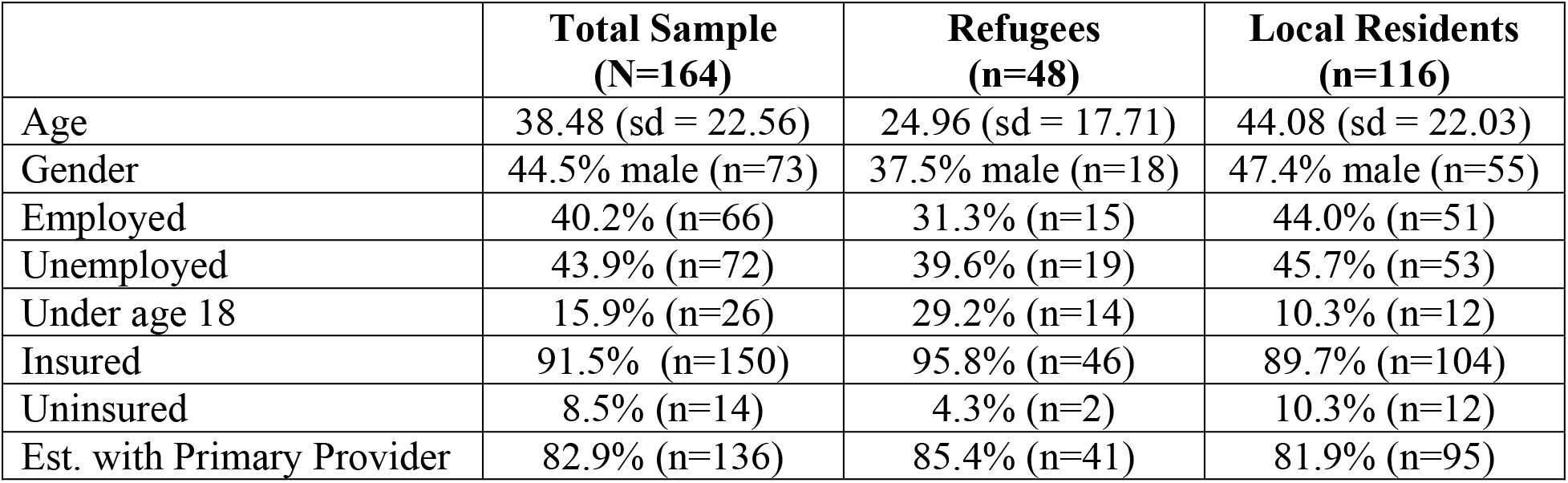
Participant Demographic Information

On average, local resident participants were significantly older than refugee participants (*t =* 5.3, p = .03). No significant between-group differences were found for other demographic variables (p > .05). Given the significant difference in mean age between refugee and local resident participants, age was included as a covariate in all subsequent analyses.

### Patient-Selected Reasons for ED Visit

The logistic regression model investigating clinical difficulties was statistically significant, X^2^(2) = 18.066, <.001. Refugee status was a significant predictor of the endorsement of clinic difficulties (p = .002). Refugees were 3.5 times more likely than local residents to report clinic difficulties as a reason for presenting to the ED. The logistic regression model investigating specialized care was statistically significant, X^2^(2) = 12.623, .002. Local resident status was a significant predictor of the endorsement of needing specialized care (p = .028). Local residents were 2.5 times more likely than refugee participants to report needing specialized care as a reason for presenting to the ED. As less than 10% of the sample reported psychosocial reasons for presenting to the ED, a logistic regression for this category was not completed. The proportion of refugees and local residents who endorsed each ED visit reason category is presented in Table 2.

**Table 2:** Proportion of Reason for ED Utilization and Use of Primary Care Physician Prior to ED Care by Group

|  | **Total Sample** | **Refugees** | **Local Residents** |
| --- | --- | --- | --- |
| Clinic Difficulties | 43.6% (71) | 68.1% (n=32) | 33.6% (n=39) |
| Specialized Care | 36.8% (n=62) | 19.1% (n=9) | 44.0% (n=51) |
| Psychosocial Reasons | 6.7% (n=11) | 6.4% (n=3) | 6.9% (n=8) |
| Called Primary Prior to ED | 46.3% (62/134) | 24.4% (n=10) | 55.9% (n=52) |

### Use of Primary Care Provider Prior to ED Visit

The majority of the sample (82.9%; n=136) reported being established with a primary care provider, with no significant differences between groups. For those who reported having a primary care provider, a logistic regression was performed to determine the effects of refugee status on the likelihood that participants would report calling their primary care provider prior to the ED visit. The logistic regression model was statistically significant, X^2^(2) = 12.843, .002. Local resident status was a significant predictor (p = .028): Local residents were 3.3 times more likely than refugees to report calling their primary care provider prior to presenting to the ED. The proportion of refugees and local residents who reported calling their primary care provider prior to their ED visit is presented in Table 2.

### Services Received During ED Visit

The logistic regression model on symptomatic care was statistically significant, X^2^(2) = 6.147, p = <.05. Refugee status was a significant predictor of receiving symptomatic care (p = .039). Refugees were 2.3 times more likely than local residents to receive symptomatic care in the ED. The logistic regression model on advanced imaging was statistically significant, X^2^(2) = 65.948, p = <.001. Both age (p <.001) and local resident status (p < .001) were significant predictors of receiving advanced imaging. Increasing age was associated with an increased likelihood to receive advanced imaging. Local residents were 24.3 times more likely to receiving advanced imaging than refugees. Refugee status was not a significant predictor of receiving specialty intervention in the ED. The proportion of refugees and local residents who received each care service type is presented in Table 3.

**Table 3:** Proportion of Care Services Received in the ED by Group

|  | **Total Sample** | **Refugees** | **Local Residents** |
| --- | --- | --- | --- |
| Symptomatic Care | 58.5% (n=96) | 72.9% (n=35) | 52.6% (n=61) |
| Specialty Intervention | 42.1% (n=69) | 27.1% (n=13) | 48.3% (n=56) |
| Advanced Imaging | 44.5% (n=73) | 4.2% (n=2) | 61.2% (n=71) |
| Admitted to Hospital | 20.7% (n=34) | 6.3% (n=3) | 26.7% (n=31) |

A logistic regression was also performed to determine the effects of refugee status on the likelihood of being admitted to the hospital. The logistic regression model was statistically significant, X^2^(2) = 26.425, p = <.001. Increasing age was associated with an increased likelihood to be admitted (p <.001). Refugee status did not significantly predict hospital admission when controlling for age (p > .05). The proportion of refugees and local residents who were admitted to the hospital is presented in Table 3.

### Discharge Diagnoses

Discharge diagnosis information by group is presented in Table 4. Four separate logistic regressions were performed to determine the effects of refugee status on the likelihood of receiving a discharge diagnosis within the following categories: infectious disease, gastrointestinal, cardiology, and musculoskeletal. All other diagnostic categories were too infrequent in the sample for meaningful analysis.

**Table 4:** Discharge Diagnostic Category by Group

|  | **Total Sample**<br>**(n=161)** | **Refugees (n=47)** | **Local Residents**<br>**(n=114)** |
| --- | --- | --- | --- |
| **Infectious Disease** | 28.0% (n=45) | 42.6% (n=20) | 21.9% (n=25) |
| **Gastrointestinal** | 16.8% (n=27) | 21.3% (n=10) | 15.8% (n=18) |
| **Cardiology** | 15.5% (n=25) | 6.4% (n=3) | 19.3% (n=22) |
| **Musculoskeletal** | 14.9% (n=24) | 10.6% (n=5) | 16.7% (n=19) |
| **Neurology** | 7.5% (n=12) | 2.1% (n=1) | 9.6% (n=11) |
| **Urology** | 6.2% (n=10) | 0 | 8.8% (n=10) |
| **OBGYN** | 5.0% (n=8) | 14.9% (n=7) | 0.9% (n=1) |
| **Pulmonology** | 4.3% (n=7) | 4.3% (n=2) | 4.4% (n=5) |
| **Endocrinology** | 2.5% (n=4) | 0 | 3.5% (n=4) |
| **ENT** | 2.5% (n=4) | 0 | 3.5% (n=4) |
| **Immunology/ONC** | 1.9% (n=3) | 2.1% (n=1) | 1.8% (n=2) |
| **Dental** | 1.9% (n=3) | 2.1% (n=1) | 1.8% (n=2) |
| **Minor Trauma** | 2.5% (n=4) | 0 | 3.5% (n=4) |
| **Psychiatry** | 1.9% (n=3) | 0 | 2.6% (n=3) |
| **Other (Genital/Urinary,**<br>**Dermatology, General Surgery,**<br>**Ophthalmology)** | 2.5% (n=4) | 4.3% (n=2) | 1.8% (n=2) |

The logistic regression model predicting infectious disease was statistically significant, X^2^(2) = 8.444, p = <.015. Age did not significantly predict an infectious disease diagnosis. Refugee status was approaching significance (p = .059) in predicting an infectious disease diagnosis at the ED when controlling for age. Refugees were 2.1 times more likely than local residents to receive an infectious disease diagnosis. Refugee status was not a significant predictor of receiving a gastrointestinal, cardiology, or musculoskeletal diagnosis (p > .05).

## DISCUSSION

Emergency Departments have traditionally served as important healthcare access points for treatment of emergent and urgent conditions in the United States. In 2015, during the study period, there were more than 143 million ED visits nationally, the highest recorded over the period of 2009-2018.^[14]^ The purpose of the current research was to identify factors influencing the decision to present to the ED among East African refugees compared to local residents in Central Minnesota, one of the largest resettlement areas of East African refugees in the United States. Further, we hoped to better understand the diagnoses and care received by refugees in the ED compared to local residents.

This study is the first to capture self-reported reasons for emergency service utilization by East African refugees. Findings showed that East African refugees were 3.5 times more likely to self-report clinic difficulties (e.g., difficulties with obtaining appointments, being available during clinic hours, and negative previous experiences at the clinic) as a reason for presenting to the ED, while local resident participants were 2.5 times more likely to report needing specialized care.

Our findings are similar to results from a study on immigrant Latinx pediatric patients who presented to the ED ^[15]^, which found that patients often presented to the ED for nonemergent issues due to parent dissatisfaction with primary care services.

Among those who reported having a PCP, local resident participants were more likely than refugees to report calling their primary care clinic prior to presenting to the ED. This finding may be a reflection of above results related to refugees acknowledging clinic difficulties as a barrier to care. Additionally, this finding may suggest difficulties with health literacy, defined as the degree to which people are able to receive and understand health information in a way that supports health decision-making.^[16]^ Existing literature shows that many refugees have low health literacy, which contributes to health disparities.^[17]^

In terms of care in the ED, refugees were 2.3 times more likely than local residents to receive symptomatic care (e.g., administration of oral or intramuscular analgesics, antiemetics, antacids, antitussive agents, steroids, bronchodilators, and anxiolytics). The majority of these interventions could be received in a primary care setting, and as such, our findings may reflect over-use of the ED by East African refugees for non-urgent conditions. This data also seems to align with previous research on asylum seekers in Switzerland, which found that 43.4% of asylum seekers presented to the ED with non-urgent medical needs, compared to only 27.9% of Swiss nationals.^[18]^ Taken together, these findings further support the need for health literacy interventions tailored to specific refugee populations, as improving refugees’ understanding of emergent-versus non-emergent medical needs may allow for more effective healthcare utilization overall.

We also investigated the types of imaging received by patients in the ED. Local residents were 24.3 times more likely to receive advanced imaging, even when controlling for age. These findings may partially reflect our finding that East African refugees were provided more symptomatic care in the ED, and as such may have had decreased need for advanced imaging compared to local residents. At the same time, the effect size for this finding suggests additional factors may be at play. Notably, only 4.2% of East African refugees in the current study received advanced imaging, in stark contrast to previous research on ED utilization of refugees. A recent Turkish study showed that 28.1% of refugees received radiologic imaging as part of their ED visit ^[19]^, and another Turkish study reported that 50% of a sample of Syrian refugees requested imaging.^[20]^ To the best of our knowledge, our study is the first to highlight differences in advanced imaging received in the ED settings by refugees compared to local residents in the United States. As such, further research on advanced imaging use in refugee populations is needed, both in comparison to other refugee communities and in comparison to local residents.

Finally, we examined discharge diagnoses received by participants after being seen by the ED provider. Refugee status was approaching significance (p = .059) in predicting an infectious disease diagnosis: refugee participants were more likely than local resident participants to receive an infectious disease diagnosis. Of note, infectious disease was the most frequent diagnosis for both local residents and refugees. Previous research in other countries suggests that rates of disease do not significantly differ between refugees and host populations.^[21]^ Given that findings from the current study were only seen at trend level, replication in larger samples in the United States is needed.

A number of limitations to the current study are worth noting. First, data was collected from a small number of the East African community in Central Minnesota, and thus, caution should be utilized when applying study results to more urban East African communities. It should also be noted that the sample size was not proportionate, which could contribute to over-representation of the experience of local residents. However, the use of bootstrapping in all analyses addressed this methodological concern, and as such, our results can be considered valid and representative of the refugee population surveyed.

We also collected data using a self-reported questionnaire written by English-speaking researchers. Even with the use of interpreters, language difficulties can occur, which could leave room for survey questions to be misinterpreted. In addition, participants might not have had a good understanding of the topic area, particularly if our findings suggest issues with health literacy in the refugee population. However, patient perspective is important in working with Somali-speaking refugees, and as such, current study findings still represent an important contribution to the literature.

Despite limitations, results suggest implications for practice in emergency medicine. Given self-reported difficulties with accessing primary care services by East African refugee participants, it may be beneficial to conceptualize the ER as an ideal place to integrate health literacy interventions for refugee populations, with the goal of improving their use of health care resources. Previous literature has shown that improvement in health literacy can lead to improved health outcomes in East African populations, as evidenced by a recent study that found higher health literacy rates led to more preventative health measures among a sample of 302 refugees from Somalia.^[22]^ Efforts to improve health literacy may include: patient education on differentiating emergencies from non-emergencies, expanding access to after-hours primary care services, and more intentional integration of diversity-sensitive and trauma-sensitive care. We hope that this data supports further exploration of health priorities for the East African refugee population, with an eye towards facilitating efforts towards equity in the public health sphere.

## Data Availability

All data produced in the present study are available upon reasonable request to the authors.

